# THE FACTOR STRUCTURE OF THE PATIENT HEALTH QUESTIONNAIRE-9 IN STROKE: A COMPARISON WITH A NON-STROKE POPULATION

**DOI:** 10.1101/2023.03.24.23287728

**Authors:** J.J. Blake, T. Munyombwe, F. Fischer, T.J Quinn, C.M. Van der Feltz-Cornelis, J.M. De Man-van Ginkel, I.S. Santos, H.J. Jeon, S. Köhler, M.T. Schram, C.D.A. Stehouwer, Jianli Wang, H.F. Levin-Aspenson, M.A. Whooley, S.E Hobfoll, S.B. Patten, A. Simning, F. Gracey, N.M. Broomfield

**Affiliations:** Department of Clinical Psychology and Psychological Therapies, University of East Anglia, Norwich, UK, NR4 7TJ; School of Medicine, University of Leeds, UK; Department of Psychosomatic Medicine, Center for Internal Medicine and Dermatology, Charité – Universitätsmedizin Berlin, corporate member of Freie Universität Berlin, Humboldt-Universität zu Berlin, and Berlin Institute of Health, Berlin, Germany; School of Cardiovascular and Metabolic Health, University of Glasgow, Glasgow, UK; Department of Health Sciences, Hull York Medical School, YBRI, University of York, UK and Institute of Health Informatics, University College London, London, UK; Nursing Science, Julius Center for Health Sciences and Primary Care, University Medical Center Utrecht, Utrecht University, Utrecht; The Netherlands & Nursing Science, Department of Gerontology and Geriatrics, Leiden University Medical Centre, Leiden, The Netherlands; Post-graduation Program in Epidemiology, Faculty of Medicine, Federal University of Pelotas, Brazil; Department of Psychiatry, Depression Center, Samsung Medical Center, Sungkyunkwan University School of Medicine, Korea; CARIM School for Cardiovascular Diseases, MHeNs School for Mental Health and Neuroscience, and Department of Internal Medicine, Maastricht University, The Netherlands; Department of Community Health & Epidemiology, Faculty of Medicine, Dalhousie University, Canada; Department of Psychology, University of North Texas, USA; Departments of Medicine, Epidemiology & Biostatistics, University of California, USA; STAR: Stress, Anxiety, and Resilience Consultants, Sandy, Utah, USA; Departments of Community Health Sciences and Psychiatry, University of Calgary, Canada; Department of Psychiatry, University of Rochester, USA

## Abstract

**Background:** There are concerns that the measurement of depression by the Patient Health Questionnaire-9 (PHQ-9), a self-report screening questionnaire, is biased by comorbid stroke sequelae. We, therefore, aimed to investigate these concerns in stroke, benchmarked against a non-stroke comparison sample, using factor analysis.

**Methods:** The secondary data sample constituted 787 stroke and 12,016 non-stroke participants, in a cross-sectional design. A subsample of 1,574 non-stroke participants was selected via propensity score matching. Dimensionality was assessed by comparing fit statistics of one-factor, two-factor, and bi-factor models. Between-group differences in factor structure were identified using measurement invariance.

**Results:** A two-factor model, consisting of somatic and cognitive-affect factors, had a superior fit to a unidimensional model (CFI = .984 versus CFI =.974, p<.001), but the high correlation between the factors indicated unidimensionality (r = .866). Configural invariance between stroke and non-stroke was supported (CFI = .983, RMSEA = .080), as were invariant thresholds (p = .092) and loadings (p = .103) for all items. Strong invariance was violated (p < .001, ΔCFI = -.003), indicating non-invariant item intercepts. Partially invariant models indicated responsibility of the tiredness and appetite intercepts, and latent depression severity was significantly overestimated in stroke, relative to the general population, using a summed score approach (Cohen’*s d=*.434).

**Conclusions:** The findings suggest that the PHQ-9 measures a single latent factor in stroke. However, the presence of non-invariant intercepts means that PHQ-9 total scores may be disproportionately influenced by fatigue in post-stroke vs. non-stroke patients and that total scores are incomparable between groups.

## INTRODUCTION

The Patient Health Questionnaire-9 (PHQ-9) is one of the most widely used depression screening tools in stroke and has demonstrated acceptable reliability, validity and classification accuracy in this population^1–3^. Despite these advantages, concerns about the applicability of the PHQ-9 in stroke remain^4^. Several items contained within the PHQ-9, such as those relating to tiredness and concentration, may also capture experiences caused by other common complications in stroke recovery, such as post-stroke fatigue^5^. Even though there are often associations between depression and, for example, post-stroke fatigue or cognitive impairment, there is also evidence that these sequelae are clinically distinct with significant variance not explained by depression^6^. Thus, loading of this unshared variance could result in unintended multidimensioanlity^4,7^. Multidimensionality, if not accounted for, can obfuscate clinical interpretation of scores and add noise to the optimal cut-off estimations if there is a general pattern of score inflation due to background physical comorbidity. This, in turn, could and cause misclassification of individuals at increased risk of depression, potentially impacting treatment received^8^.

Two recent publications have provided partial support for the unidimensionality of the PHQ-9 in stroke and robustness to extraneous sequelae, with a preliminary indication that the PHQ-9 may trend towards insufficient unidimensionality with increased time since stroke^9,10^. However, measurement accuracy can be compromised in more subtle ways, such as affecting item category thresholds or intercepts^8^. Differences in these parameters between populations could invalidate statistical comparisons. For example, unequal intercepts between populations can bias depression estimates using the traditional summed score approach, and potentially lead to a greater likelihood of type I and II errors^8^.

It is therefore important that the dimensionality of the PHQ-9 in stroke is further appraised and that the factor structure of the PHQ-9 in stroke is compared with other populations to understand population-level effects. We address the following aims in this study:

1. To assess the dimensionality of the PHQ-9 in a stroke population
2. To identify differences in factor structure that might be attributable to stroke via measurement invariance comparison of non-stroke samples
3. To explore the significance of any non-invariance of item thresholds, loadings, intercepts, or residual variances between groups

## METHODS

### Design

We conducted a cross-sectional study using secondary data. We identified studies with eligible data from primary studies included in a previous individual participant meta-analysis^3^ and requested data from the primary study authors. Studies spanned multiple nations and languages (Table 1).

**Table 1.** Demographic data, stratified by cluster

| Study | n | % female | Age (SD) | Country | Language | Population description/setting | Mean PHQ-9 (SD) | Approximate time since stroke (months) |
| --- | --- | --- | --- | --- | --- | --- | --- | --- |
| **Stroke** |  |  |  |  |  |  |  |  |
| De Man-Van Ginkel (2012) | 382 | 45.8 | 69.2 (14.5) | Netherlands | Dutch | Acute stroke patients | 6.6 (5.6) | 0.3 – 2.6 (M: 1.58) |
| Prisnie (2016) | 114 | 56.1 | 59.6 (15.5) | Canada | English | Outpatient community stroke | 4.8 (5.2) | 2.2 – 10.6 (M: 3.6) |
| Quinn (unpublished) | 135 | 47.7 | 68.4 (12.7) | UK | English | Inpatient acute stroke | 6.7 (6.3) | 0 – 0.45 |
| Simning (2018) | 21 | 47.6 | 70.0 (6.5) | US | English | Older adults in public housing | 4.7 (4.4) | Unavailable |
| Thombs (2008) | 144 | 16.7 | 69.7 (10.1) | US | English | People with Coronary Artery Disease in the community | 5.8 (5.6) | Unavailable |
| **Total** | 796 | 42.3 | 67.8 (13.9) |  |  |  | 6.1 (5.6) |  |
| **Non-stroke Comparison Samples** |  |  |  |  |  |  |  |  |
| Kim (2017) | 3071 | 56.6 | 38.8 (12.2) | South Korea | Korean | Randomly selected adults, via S. Korean census | 2.2 (2.2) |  |
| Liu (2015) | 4182 | 55.0 | 44.7 (10.0) | Canada | English | Working population | 3.2 (4.0) |  |
| Janssen (2016) | 3502 | 55.3 | 59.5 (8.6) | Netherlands | Dutch | Data from The Maastricht Study: a population-based cohort study, recruited from multiple sites | 2.7 (3.2) |  |
| Levin-Aspenson (2018) | 408 | 68.6 | 45.0 (13.4) | US | English | General population community-based adults | 6.5 (6.6) |  |
| Santos (2013) | 447 | 57.3 | 43.8 (15.1) | Brazil | Portuguese | General population via random household sampling | 5.0 (5.1) |  |
| Simning (2012) | 169 | 59.2 | 67.3 (6.6) | US | English | Older adults in public housing | 5.1 (4.3) |  |
| Volker (2016) | 93 | 50.5 | 46.4 (10.9) | Netherlands | Dutch | Employees on sickness leave in an occupational health setting | 8.5 (7.6) |  |
| Hobfoll (2011) | 144 | 57.7 | 41.6 (15.2) | Israel | Hebrew and Arabic | Jewish and Palestinian residents of Jerusalem exposed to war | 5.9 (5.9) |  |
| **Total** | 12016 | 56.0 | 47.8 (13.5) |  |  |  | 3.1 (4.0) |  |

### Participants

We sampled five studies with stroke patients ^11–15^, covering inpatient and community settings. Individual stroke characteristic data, such time since stroke, were not available. General post-stroke time points, where available, are outlined in Table 1. The comparison group was sampled from eight studies^16–23^. The comparison group were recruited from a variety of settings, generally free of significant health morbidities. Most sampled the general population^18–20,23^, but three of the smaller samples were recruited from specific groups or contexts^16,21,22^.

Exclusion and inclusion criteria are specified in the source studies. We required that stroke source studies confirmed the presence of a non-transient stroke. Comparison group studies were excluded if they focused on any specific long-term health condition, though the existence of such conditions amongst individual participants, was permitted. Stroke status was not recorded in most comparison group samples, so stroke-free status could not be guaranteed. However, this was only anticipated to make up a minority of cases based on published incident rates^24^.

### Measure

Each PHQ-9 item corresponds to one of the nine DSM-IV depression criteria^25^ and is scored on a four-category Likert scale, relating to the frequency of the symptom experienced in the past two weeks. Item scores are added to calculate a measure total, with a maximum score of 27. An optimal cut-off of ≥10 is generally suggested in the literature for stroke and the general population^1,3^.

### Analysis

Propensity score matching was used to select a sample from the non-stroke dataset that was more demographically aligned to the stroke sample, using the MatchIt package in R^26^. Age, sex, country, and PHQ-9 total score were entered for matching. PHQ-9 total score was matched to align crude depression status as additional population control. Missing data were excluded listwise.

#### Assessment of dimensionality

Confirmatory Factor Analysis (CFA) was modelled in R using Lavaan^27^ and SEMTools^28^. A single-factor CFA model, representing generalised depression, was evaluated alongside two variations of a two-factor model and a bi-factor model. The two-factor models each consisted of a somatic and cognitive-affective factor but differed concerning which factor items seven, relating to trouble concentrating, and eight, relating to feeling slowed down, were specified because of inconsistencies in the literature about the optimal specification^4,7^.

The bi-factor model consisted of a general factor and two specific factors, cognitive-affective and somatic^29^. The factor location of items seven and eight in the bifactor model was determined by examining which of the two-factor models had a superior fit. Bifactor models can indicate dimensionality through comparisons of correlations of bifactor global factor scores with one-factor model scores. If multidimensionality were present, the global depression factor in the bifactor model would lose significant variance to the specific, uncorrelated, factors because a large proportion of somatic item variance would not be due to depression and thus load to the specific somatic factor instead of the global depression factor. This would result in a comparatively weak correlation between bifactor global and one-factor model scores.

Each CFA model was fitted using Diagonally Weighted Least Squares (DWLS) estimation, which is suitable for ordinal-level data and cases where multivariate normality is violated. Robust equivalents of Root Mean Square Error of Approximation (RMSEA; values of <0.08 interpreted as acceptable fit) and Comparative Fit Index (CFI; >0.96 interpreted as acceptable fit) were used to evaluate model fit^30^. Fit statistics of the single were compared to identify whether one or two latent factors best described the covariances in item responses in the stroke sample.

#### Measurement invariance

The stroke and comparison groups were assessed for measurement invariance, using established procedures for ordinal measures, to evaluate possible differences in factor structure.^31^ This involves equality testing of four parameters between groups: item thresholds, loadings, intercepts, and residual variances, sequentially.^32^ Item thresholds are the point on the item’s latent continuum in which people, on average, start to endorse each ordinal category, loadings are the degree of association of an item with its respective factor, item intercepts are the expected value of the item if the latent factor is set to 0, and residual variances are variances that is not accounted for by the latent factors. If the groups do not significantly differ in these parameters, the models are considered to be invariant and scores therefore comparable^8^. Invariance of thresholds is henceforth referred to as ‘threshold invariance’, invariance of loadings as ‘metric invariance’, invariance of item intercepts as ‘scalar invariance’, and invariance of item residuals as ‘full invariance’^33^. In cases of non-invariance, a change in the direction of poorer fit would be expected because the model specifies equivalences that are not reflected in the data.

As per this methodology, a baseline multi-group CFA model, referred to as a configural model, was fit. Progressive equality restraints were sequentially applied and tested against the previous model. Changes to fit after each stage were examined via two methods: a one-way ANOVA significance test in chi-square fit statistics, using the Satorra (2000) method,^34^ and by inspecting changes to the CFI and RMSEA. Increases in *X*^2^ and RMSEA values and decreases in CFI values are indicative of reduced model fit. Changes in CFI >.001 are commonly interpreted as an indicator of measurement non-invariance^35^.

Chi-square difference tests are sample-size dependent and find significant differences among small non-meaningful effects in large samples^32^. However, caution is advised with interpreting changes to CFI and RMSEA when using ordered data and DWLS estimation^36^. A pragmatic approach was, therefore, adopted; non-significant changes to chi-square values were assumed to be robust indicators of invariance, given the large samples. In cases where a significant p-value of chi-square difference was observed, a detailed exploration of invariance violation was explored to identify the meaningfulness of the observed differences.

#### Sample size

A sample size of 300-500 for CFA modelling has demonstrated robustness to low communalities and loadings, with a minimum of 200^37^. Each stroke sample was insufficient in size to be modelled separately, so these clusters were combined. The sample size in each group was therefore sufficient for robust parameter estimation. Statistically, accounting for clustering was impractical because of the number of clusters, the small size of each cluster, and the limitations of current software capabilities.

#### Ethics

This study was approved by the University of East Anglia Faculty of Medicine and Health Research Ethics Committee (UEA FMH REC) on 11^th^ August 2021 (approval number: 2020/21-046). Participation consent had been provided to primary authors.

## RESULTS

### Data processing and matching

Eight cases of missing data were removed from the stroke sample and five from the non-stroke sample before propensity matching, constituting just .11% of the original dataset. As suspected, substantial differences were found between stroke and non-stroke comparison samples in sex, nationality, language, age, and PHQ-9 total score (Table 2). Propensity matching was, therefore, necessary to reduce these differences. A 2:1 ratio sample was selected for analysis.

**Table 2.** Demographic overview of samples before and after matching

|  |  | Pre-matched<br>Comparison<br>Group<br>(n = 12,011) | Stroke<br>(n = 787) | Diff | Test<br>statistic | p | Effect<br>statistic | Effect size<br>descriptor |
| --- | --- | --- | --- | --- | --- | --- | --- | --- |
| %<br>Female | | 56.1% | 42.3% | 13.8% | 56.92<br>($X^2$) | <.001 | .067 ($\phi$) | Negligible |
| Age | | 47.8 (SD 13.5) | 67.8 (SD 13.9) | -20.1 | -40.26<br>(t) | < .001 | 1.46 ($d$) | Large |
| Country | UK | 0.0% | 16.0% | -16.0% | 2669.64<br>($X^2$) | < .001 | .46 ($V$) | Large |
|  | US | 4.8% | 21.0% | -16.2% |  |  |  |  |
|  | Canada | 34.8% | 14.5% | 20.3% |  |  |  |  |
|  | Netherlands | 29.9% | 48.5% | -28.5% |  |  |  |  |
|  | Brazil | 3.7% | 0.0% | 3.7% |  |  |  |  |
|  | Israel | 1.2% | 0.0% | 1.2% |  |  |  |  |
|  | South Korea | 25.6% | 0.0% | 25.6% |  |  |  |  |
|  | **Total** | 100% | 100% |  |  |  |  |  |
| Language | English | 39.6% | 51.5% | -11.9% | 347.84<br>($X^2$) | < .001 | .17 ($V$) | Small |
|  | Dutch | 29.9% | 48.5% | -28.5% |  |  |  |  |
|  | Portuguese | 3.7% | 0.0% | 3.7% |  |  |  |  |
|  | Hebrew | 0.6% | 0.0% | 0.6% |  |  |  |  |
|  | Arabic | 0.6% | 0.0% | 0.6% |  |  |  |  |
|  | Korean | 25.6% | 0.0% | 25.6% |  |  |  |  |
|  | **Total** | 100% | 100% |  |  |  |  |  |
| PHQ-9<br>total | | 3.1 (4.0) | 6.1 (5.6) | -3.0 | -14.9 (t) | <.001 | .62 ($d$) | Medium |
|  |  | Matched<br>Comparison<br>Group<br>(n = 1574) | Stroke<br>(n = 787) | Diff | Test<br>statistic | p | Effect<br>statistic | Effect size<br>descriptor |
| %<br>Female | | 41.4% | 42.3% | -0.9% | .17 ($X^2$) | .679 | .01 ($\phi$) | Non-significant |
| Age | | 61.8 (SD 11.3) | 67.8 (SD 13.9) | -6.04 | -10.56<br>(t) | < .001 | .48 ($d$) | Medium |
| Country | UK | 0.0% | 16.0% | -16.0% | 349.37<br>($X^2$) | < .001 | .39 ($V$) | Medium |
|  | US | 17.1% | 21.0% | -3.9% |  |  |  |  |
|  | Canada | 30.3% | 14.5% | 15.8% |  |  |  |  |
|  | Netherlands | 47.5% | 48.5% | -1.0% |  |  |  |  |
|  | Brazil | 2.8% | 0.0% | 2.8% |  |  |  |  |
|  | Israel | 1.2% | 0.0% | 1.2% |  |  |  |  |
|  | South Korea | 1.1% | 0.0% | 1.1% |  |  |  |  |
|  | **Total** | 100% | 100% |  |  |  |  |  |
| **Language** | English | 47.4% | 51.5% | -4.1% | 42.41<br>($X^2$) | < .001 | .13 (*V*) | Small |
|  | Dutch | 47.5% | 48.5% | -1.0% |  |  |  |  |
|  | Portuguese | 2.8% | 0.0% | 2.8% |  |  |  |  |
|  | Hebrew | 0.7% | 0.0% | 0.7% |  |  |  |  |
|  | Arabic | 0.5% | 0.0% | 0.5% |  |  |  |  |
|  | Korean | 1.1% | 0.0% | 1.1% |  |  |  |  |
|  | **Total** | 100% | 100% |  |  |  |  |  |
| **PHQ-9 total** |  | 5.9 (6.0) | 6.1 (5.6) | .24 | -.95 (t) | .343 | .04 (*d*) | Non-significant |

The final sample, after matching and removal of missing data, consisted of 787 stroke and 1574 comparison participants. Demographic details of the stroke group and propensity-matched comparison group, including significance tests of demographic differences, are summarised in Table 2. Compared with the pre-matched sample, the demographic differences in the matched sample were substantially reduced for each variable, with non-significant gender and PHQ-9 total score differences. Significant differences remained for nationality, language, and age, with medium to large effect sizes. Despite findings of significant differences in nationality, most participants in both groups were from western developed nations (94.9% in non-stroke and 100% in stroke) and may, therefore, represent similar cultural backgrounds.

### The Dimensionality of the PHQ-9 in Stroke

Fit statistics for the one-factor model, each two-factor model, and the bifactor model are summarised in Table 3 for stroke and non-stroke groups. All models had sufficient fit to the data, based on CFI values >.96 in both groups. All stroke group models had a sufficient RMSEA fit of <.08, but the non-stroke one-factor model did not.

**Table 3.** Specification and fit statistics of stroke CFA models

| Model | Factors | Model specification<br>(numbers denote items) | Model<br>parameters | Stroke group |  |  |  | Comparison group |  |  |  |
| --- | --- | --- | --- | --- | --- | --- | --- | --- | --- | --- | --- |
|  |  |  |  | X2 (df) | Robust<br>CFI | Robust<br>RMSEA | Factor<br>correlation | X2<br>(df) | Robust<br>CFI | Robust<br>RMSEA | Factor<br>correlation |
| One-factor | 1 | All items onto a single factor | 36 | 129.04<br>(27) | .974 | .069 | - | 355.9<br>(27) | .982 | .088 | - |
| Two-factor<br>A | 2 | Cognitive/affective: 1,2,6 9<br>Somatic: 3, 4, 5, 7, 8 | 37 | 88.61<br>(26) | .984 | .055 | .866 | 212.1<br>(26) | .990 | .067 | .910 |
| Two-factor<br>B | 2 | Cognitive/affective: 1,2,6,<br>7, 8, 9<br>Somatic: 3, 4, 5 | 37 | 108.27<br>(26) | .979 | .063 | .865 | 252.1<br>(26) | .988 | .074 | .908 |
| Bifactor | 3 | Global depression: all items<br>Cognitive/affective: 1,2,6 9<br>Somatic: 3, 4, 5, 7, 8<br>Factor variances are freely estimated. Correlations between factors set to 0 | 45 | 39.36<br>(18) | .995 | .039 | Set to 0 | 121.8<br>(18) | .994 | .061 | Set to 0 |

$\chi^2$ = Chi-squared, df = degrees of freedom, CFI = Comparative Fit Index, RMSEA = Root Mean Square Error of Approximation. Item 1=little interest, item 2=down/depressed/hopeless, item 3=sleep problems, item 4=tiredness, item 5=appetite, item 6=feeling bad about oneself, item 7=trouble concentrating, item 8= moving slowly, and item 9=suicidality.

Of the two alternate forms of the two-factor model, the model that specified problems with concentration and moving slowly in the somatic factor, two-factor A, had a superior fit in both groups. This suggests that items 7 and 8 covary more strongly with the somatic items 3, 4, and 5, relating to sleep, tiredness, and appetite, respectively. The stroke two-factor A model had a superior fit to the stroke one-factor model, *X*^*2*^ diff = 24.83, df diff = 1, p <.001. However, a high correlation (r =.865) was observed between factors in stroke, which is indicative of practical unidimensionality.

To further assess dimensionality in the stroke group, global depression factor scores were calculated from the bifactor model and plotted against latent depression scores from the one-factor model. The correlation between latent depression scores was .99 in the stroke group, indicating substantial shared variance in factor-derived scores and practical unidimensionality.

### Measurement invariance

Findings of measurement invariance testing are summarised in Table 4. The unidimensional configural model possessed sufficient fit (CFI > .96, RMSEA = .080), indicating a similar factor structure between the groups.

**Table 4.** Unstandardised *X*^2^ fit statistics of multi-group CFA at each level of constraint *X*^*2*^*= Chi-squared, df = degrees of freedom, CFI = Comparative Fit Index, RMSEA = Root Mean Square Error of Approximation*, ***Δ=*** *change in the associated fit statistic*, the p-value corresponds to the significance of the change of chi-square of model fit.

| Model | $X^2$ (df) | CFI scaled | RMSEA scaled | $\Delta X^2$ ($\Delta df$) | p | $\Delta CFI$ | $\Delta RMSEA$ |
| --- | --- | --- | --- | --- | --- | --- | --- |
| Configural model | 268.6 (54) | .983 | .080 |  |  |  |  |
| Constrained thresholds | 272.3 (63) | .981 | .077 | 15.0 (9) | .092 | -.001 | -.003 |
| Constrained loadings | 284.2 (71) | .982 | .072 | 13.2 (8) | .103 | .000 | -.005 |
| Constrained intercepts | 403.8 (79) | .979 | .074 | 57.8 (8) | <.001* | -.003 | .001 |

The constraining of item thresholds and loadings to be equal between groups led to non-significant increases in unstandardised chi-square statistics and only marginal changes to CFI and RMSEA values, indicating equal thresholds and loadings. A significant reduction of model fit was observed after the constraint of item intercepts, as indicated by the p-value for the chi-square difference.

To identify the intercepts responsible for the violation of scalar invariance, nine partially invariant CFA models were specified, with item intercept constraints released one at a time. Substantial between-group differences for item 4, relating to tiredness, and 5, relating to appetite, were found. The tiredness intercept was substantially greater in the stroke group, with a relative difference of .446, and the appetite intercept was significantly greater in the matched comparison group, with a relative difference of .346. The average absolute magnitude of intercept differences of the remaining items was just .07. These findings indicate that tiredness scores are higher in the stroke group and appetite disruption scores are higher in the comparison group when controlling for latent depression severity.

A partially invariant model, which estimated the intercepts of items 4 and 5 freely while maintaining constraints for the remaining items, was specified to confirm that between-groups differences in intercepts for items 4 and 5 were responsible for failed scalar invariance. The fit of the partial model (*X*^*2*^*=*296.2, CFI= .985, RMSEA = .064) was statistically compared to the constrained loadings model (*X*^*2*^ *=* 284.2, CFI = .982, RMSEA = .072), with no significant reduction in model fit observed, *X*^*2*^ diff = 7.69, df diff = 6, p = .262. This finding confirms the responsibility of items 4 and 5 for failed scalar invariance.

To identify the meaningfulness of the scalar invariance violation, the effect sizes of between-group comparisons using a traditional summed score approach and using model-derived scores were compared (see Table 5). The stroke and non-stroke groups did not significantly differ when using the traditional summed score approach, because of group matching. However, model-implied depression scores indicated significantly greater latent depression in the non-stroke group. The observed effect size was .117 larger for the partially invariant model than for the fully invariant model, which suggests that falsely assumed scalar invariance results in moderate underestimation of between-groups differences.

**Table 5.** Between-groups differences in depression scores, derived from a sum score approach and model-derived latent factor estimations for fully and partially invariant models *Negative signs indicate higher estimated depression in the stroke group, and positive signs indicate higher depression in the non-stroke group*.

| Scoring approach | Stroke<br>M | Non-stroke<br>M | t/z statistic | p | Effect size<br>(Cohen's d) | Descriptor |
| --- | --- | --- | --- | --- | --- | --- |
| Sum score | 6.141 | 5.902 | -0.94 (t) | .255 | -.045 | Non-significant |
| Fully invariant model | 0 | .272 | 4.80 (z) | < .000 | .272 | Small |
| Partially invariant<br>model | 0 | .389 | 5.91 (z) | <.000 | .389 | Small |

The effect size of between-groups comparisons of depression scores derived from the partially invariant model was .434 larger than that observed when using the summed score method, which is a difference that equates to a small-to-medium effect. The effect of unequal intercepts is, therefore, substantial, which indicates that depression scores, using a summed score approach, are incomparable between groups.

## DISCUSSION

This study aimed to assess the dimensionality of the PHQ-9 in stroke and to identify possible differences in factor structure to those in the wider population. Despite the two-factor models demonstrating a better fit than the one-factor model, we found evidence of unidimensionality in stroke. This finding contributes further evidence for PHQ-9 in favour of good psychometric performance in stroke^1^ and general robustness to violations of unidimensionality.^10^

The two groups were invariant for thresholds and factor loadings. This implies that the factor correlations of items are broadly equivalent between groups and that the thresholds in which patients move to endorse a higher response category occur at approximately equal points on the item’s latent continuum. Differences in intercepts were, however, observed; specifically, a large positive intercept of tiredness was observed in the stroke group and a large positive intercept of appetite disruption in the comparison group. This implies that stroke patients are more likely to positively endorse the tiredness item, and non-stroke participants the appetite item when latent depression is held constant.

The violation of equal intercepts was clinically significant. A small-to-medium between-groups difference in latent depression was obscured by the inequalities in intercepts, with the tiredness intercept significantly inflating PHQ-9 total scores in stroke. Despite similar summed total scores between propensity-matched samples, latent depression estimates from the partially invariant model suggest that the non-stroke sample was, on average, more depressed. This confounds the between-group comparability of scores when using a summed score approach and could result in type I and II errors in research. Such bias from the tiredness item might lead to an overdetection of individuals at increased risk of depression in those with post-stroke fatigue and an underdetection in those without post-stroke fatigue, which may have introduced noise and biased estimates of optimal cut-offs in stroke.^1^ Indeed, the potential bias of physical comorbidities and the effect on optimal cut-off points has been demonstrated in patients with diabetes, suggesting that there may be variation between post-stroke fatigue subgroups.^17,38^

A strength of this study is that it is one of the first to compare the factor structure of the PHQ-9 between stroke and non-stroke comparison groups. We have provided additional confirmation of the general robustness of the measure’s overall dimensionality to physical health problems and other stroke sequelae.^9,10^ The finding of noninvariant intercepts is also of importance because it suggests that comparisons of PHQ-9 scores between stroke and other groups might be invalid.

Despite these strengths, several limitations exist that may impact generalisability. First, it was difficult to statistically account for nested data because this would have required multiple measurement invariance calculations between groups and because lavaan has limited functionality for completing measurement invariance on nested data. Unaccounted clustering can lead to biased parameter estimations and standard errors because of the presence of item covariations within clusters that are not accounted for by the latent variable.^39^ An additional limitation of this research is the imperfect demographic matching. Despite the substantial initial sample of non-stroke comparison participants, the differences in demographics were too great for perfect matching. Finally, a minority of the comparison sample may have suffered strokes, which may have marginally increased sample error and reduced the magnitude of observed differences.

Data on the amount of time elapsed since the index stroke event were not available. As such, the stroke participants were likely to have a substantial variance in their position in stroke recovery. Time since the stroke event is important because a recent publication has demonstrated weak invariance as a factor of time since stroke^9^ and because significant improvement in physical functioning can be observed up to one-year post-stroke, as well as cognitive decline because of emerging dementias.^40^

The finding of unidimensionality suggests that clinicians can continue using the PHQ-9 in stroke practice. Clinicians should be mindful that stroke patients will report higher baseline tiredness on item 4 of the PHQ-9, and that those with significant post-stroke fatigue may have inflated total scores compared to those who do not. Caution in the interpretation of patients near the cut-off point is, therefore, advised until future research clarifies this concern. Researchers aiming to compare scores between stroke and non-stroke should consider estimating latent depression via modelling. Removal of items 4 and 5 would also address the issue of unequal intercepts but reduce content validity.

Several avenues for future research emerge from the current study. A mixed two-factor approach to measurement invariance methodology, whereby group-level and longitudinal-level measurement invariance are simultaneously investigated would promote a greater understanding of the longitudinal changes to factor structure associated with stroke recovery.^9^ The causes of the differences in intercepts should be investigated in more detail. It is also important that measurement invariance, diagnostic accuracy, and differences in optimal cut-off points are assessed between people with post-stroke fatigue and those without, because of concerns about the tiredness item.

## Data Availability

Secondary data were acquired under specific authorisation for the primary author (data usage agreements). Thus, we do not own the data, and any request for data would first need to be approved by all co-authors, and this may require additional data protection approval.

## Acknowledgements

we firmly thank Prof. Thombs, Dr Levis, Dr Benedetti, and Sheryl Sun of the DEPRESSD collaboration (McGill University) for their support in reviewing the study protocol and contacting primary authors to obtain the data used in this study.

## Data access

data are held by the corresponding author, who takes responsibility for its integrity and analysis

## REFERENCES

1. Burton L-JJ, Tyson S. Screening for mood disorders after stroke: A systematic review of psychometric properties and clinical utility. Psychological Medicine. 2015;45(1):29–49.

2. Kroenke K, Spitzer RL, Williams JBW. The PHQ-9: Validity of a brief depression severity measure. Journal of General Internal Medicine. 2001;16(9):606–613.

3. Negeri ZF, Levis B, Sun Y, He C, Krishnan A, Wu Y, Bhandari PM, Neupane D, Brehaut E, Benedetti A, Thombs BD. Accuracy of the Patient Health Questionnaire-9 for screening to detect major depression: updated systematic review and individual participant data meta-analysis. BMJ. 2021;375.

4. Chilcot J, Rayner L, Lee W, Price A, Goodwin L, Monroe B, Sykes N, Hansford P, Hotopf M. The factor structure of the PHQ-9 in palliative care. Journal of Psychosomatic Research. 2013;75(1):60–64.

5. Langhorne P, Stott DJ, Robertson L, MacDonald J, Jones L, McAlpine C, Dick F, Taylor GS, Murray G. Medical Complications After Stroke. Stroke. 2000;31(6):1223–1229.

6. Mitchell AJ, Sheth B, Gill J, Yadegarfar M, Stubbs B, Yadegarfar M, Meader N. Prevalence and predictors of post-stroke mood disorders: A meta-analysis and meta-regression of depression, anxiety and adjustment disorder. General Hospital Psychiatry. 2017;47(October 2015):48–60.

7. Krause JS, Reed KS, McArdle JJ. Factor structure and predictive validity of somatic and nonsomatic symptoms from the patient health questionnaire-9: A longitudinal study after spinal cord injury. Archives of Physical Medicine and Rehabilitation. 2010;91(8):1218–1224.

8. Kim ES, Yoon M. Testing Measurement Invariance: A Comparison of Multiple-Group Categorical CFA and IRT. http://dx.doi.org.uea.idm.oclc.org/10.1080/10705511.2011.557337. 2011;18(2):212–228.

9. Dong L, Williams LS, Briceno E, Morgenstern LB, Lisabeth LD. Longitudinal assessment of depression during the first year after stroke: Dimensionality and measurement invariance. Journal of Psychosomatic Research. 2022;153:110689.

10. Katzan IL, Lapin B, Griffith S, Jehi L, Fernandez H, Pioro E, Tepper S, Crane PK. Somatic symptoms have negligible impact on Patient Health Questionnaire-9 depression scale scores in neurological patients. European Journal of Neurology. 2021;28(6):1812–1819.

11. de Man-van Ginkel Jm, Hafsteinsdóttir T, Lindeman E, Burger H, Grobbee D, Schuurmans M, JM de MG, Hafsteinsdóttir T, Lindeman E, Burger H, Grobbee D, Schuurmans M, de Man-van Ginkel Jm, Hafsteinsdóttir T, Lindeman E, et al. An efficient way to detect poststroke depression by subsequent administration of a 9-item and a 2-item Patient Health Questionnaire. Stroke (00392499). 2012;43(3):854–856.

12. Prisnie JC, Fiest KM, Coutts SB, Patten SB, Atta CAM, Blaikie L, Bulloch AGM, Demchuk A, Hill MD, Smith EE, Jetté N. Validating screening tools for depression in stroke and transient ischemic attack patients. International Journal of Psychiatry in Medicine. 2016;51(3):262–277.

13. Simning A, Seplaki CL, Conwell Y. The association of a heart attack or stroke with depressive symptoms stratified by the presence of a close social contact: findings from the National Health and Aging Trends Study Cohort. International journal of geriatric psychiatry. 2018;33(1):96–103.

14. Thombs BD, Ziegelstein RC, Whooley MA. Optimizing detection of major depression among patients with coronary artery disease using the patient health questionnaire: data from the heart and soul study. Journal of general internal medicine. 2008;23(12):2014–2017.

15. Quinn TJ, Taylor-Rowan M, Elliott E, Drozdowska B, McMahon D, Broomfield NM, Barber M, MacLeod MJ, Cvoro V, Byrne A, Ross S, Crow J, Slade P, Dawson J, Langhorne P. Research protocol – Assessing Post-Stroke Psychology Longitudinal Evaluation (APPLE) study: A prospective cohort study in stroke. Cerebral Circulation - Cognition and Behavior. 2022;3:100042.

16. Hobfoll SE, Canetti D, Hall BJ, Brom D, Palmieri PA, Johnson RJ, Pat-Horenczyk R, Galea S. Are Community Studies of Psychological Trauma’s Impact Accurate? A Study Among Jews and Palestinians. Psychological Assessment. 2011;23(3):599–605.

17. Janssen EPCJ, Köhler S, Stehouwer CDA, Schaper NC, Dagnelie PC, Sep SJS, Henry RMA, van der Kallen Cjh, Verhey FR, Schram MT. The Patient Health Questionnaire-9 as a Screening Tool for Depression in Individuals with Type 2 Diabetes Mellitus: The Maastricht Study. Journal of the American Geriatrics Society. 2016;64(11):e201–e206.

18. Levin-Aspenson HF, Watson D. Mode of administration effects in psychopathology assessment: Analyses of gender, age, and education differences in self-rated versus interview-based depression. Psychological Assessment. 2018;30(3):287–295.

19. Liu Y, Wang JL. Validity of the Patient Health Questionnaire-9 for DSM-IV major depressive disorder in a sample of Canadian working population. Journal of Affective Disorders. 2015;187:122–126.

20. Santos IS, Tavares BF, Munhoz TN, de Almeida Lsp, da Silva Ntb, Tams BD, Patella AM, Matijasevich A. Sensibilidade e especificidade do Patient Health Questionnaire-9 (PHQ-9) entre adultos da população geral. Cadernos de Saude Publica. 2013;29(8):1533–1543.

21. Simning A, Van Wijngaarden E, Fisher SG, Richardson TM, Conwell Y. Mental healthcare need and service utilization in older adults living in public housing. The American journal of geriatric psychiatry : official journal of the American Association for Geriatric Psychiatry. 2012;20(5):441–451.

22. Volker D, Zijlstra-Vlasveld MC, Brouwers EPM, Homans WA, Emons WHM, van der Feltz-Cornelis Cm. Validation of the Patient Health Questionnaire-9 for Major Depressive Disorder in the Occupational Health Setting. Journal of occupational rehabilitation. 2016;26(2):237–244.

23. Kim DJ, Kim K, Lee HW, Hong JP, Cho MJ, Fava M, Mischoulon D, Heo JY, Jeon HJ. Internet Game Addiction, Depression, and Escape from Negative Emotions in Adulthood: A Nationwide Community Sample of Korea. Journal of Nervous and Mental Disease. 2017;205(7):568–573.

24. Hollander M, Koudstaal PJ, Bots ML, Grobbee DE, Hofman A, Breteler MMB. Incidence, risk, and case fatality of first ever stroke in the elderly population. The Rotterdam Study. Journal of Neurology, Neurosurgery & Psychiatry. 2003;74(3):317–321.

25. Bell CC. DSM-IV: Diagnostic and Statistical Manual of Mental Disorders. JAMA: The Journal of the American Medical Association. 1994;272(10):828.

26. Ho DE, Imai K, King G, Stuart EA. MatchIt: Nonparametric preprocessing for parametric causal inference. Journal of Statistical Software. 2011;42(8):1–28.

27. Rosseel Y. Lavaan: An R package for structural equation modeling. Journal of Statistical Software. 2012;48.

28. semTools Contributors. semTools: Useful tools for structural equation modeling. R package version 0.4-14. 2016.

29. Fischer F, Levis B, Falk C, Sun Y, Ioannidis JPA, Cuijpers P, Shrier I, Benedetti A, Thombs BD. Comparison of different scoring methods based on latent variable models of the PHQ-9: an individual participant data meta-analysis. Psychological Medicine. 2021:1–12.

30. Li CH. The performance of ML, DWLS, and ULS estimation with robust corrections in structural equation models with ordinal variables. Psychological Methods. 2016;21(3):369–387.

31. Wu H, Estabrook R. Identification of Confirmatory Factor Analysis Models of Different Levels of Invariance for Ordered Categorical Outcomes. Psychometrika. 2016;81(4):1014.

32. Fischer F, Gibbons C, Coste J, Valderas JM, Rose M, Leplège A. Measurement invariance and general population reference values of the PROMIS Profile 29 in the UK, France, and Germany. Quality of Life Research. 2018;27(4):999–1014.

33. Putnick DL, Bornstein MH. Measurement Invariance Conventions and Reporting: The State of the Art and Future Directions for Psychological Research. Developmental review : DR. 2016;41:71.

34. Satorra A. Scaled and Adjusted Restricted Tests in Multi-Sample Analysis of Moment Structures. 2000:233–247.

35. Khademi A, Wells CS, Oliveri ME. Examining Appropriacy of CFI/TLI Cutoff Value in Multiple-group CFA Test of Measurement Invariance to Enhance Accuracy of Test Score Interpretation. 2021.

36. Sass DA, Schmitt TA, Marsh HW. Evaluating Model Fit With Ordered Categorical Data Within a Measurement Invariance Framework: A Comparison of Estimators. Structural Equation Modeling. 2014;21(2):167–180.

37. MacCallum RC, Widaman KF, Zhang S, Hong S. Sample size in factor analysis. Psychological Methods. 1999;4(1):84–99.

38. Van Steenbergen-Weijenburg KM, De Vroege L, Ploeger RR, Brals JW, Vloedbeld MG, Veneman TF, Hakkaart-Van Roijen L, Rutten FF, Beekman AT, Van Der Feltz-Cornelis Cm. Validation of the PHQ-9 as a screening instrument for depression in diabetes patients in specialized outpatient clinics. BMC health services research. 2010;10.

39. Dyer NG, Hanges PJ, Hall RJ. Applying multilevel confirmatory factor analysis techniques to the study of leadership. The Leadership Quarterly. 2005;16(1):149–167.

40. McHutchison CA, Cvoro V, Makin S, Chappell FM, Shuler K, Wardlaw JM. Functional, cognitive and physical outcomes 3 years after minor lacunar or cortical ischaemic stroke. Journal of Neurology, Neurosurgery & Psychiatry. 2019;90(4):436–443.

